# Research Protocol for a Randomised Controlled Trial Comparing the Outcome of the 3D-Printed Patient-Specific-Instrument Assisted Lapidus Fusion vs Conventional Lapidus Fusion for Surgical Correction of Hallux Valgus Deformity

**DOI:** 10.1101/2023.10.05.23296589

**Authors:** Samuel Ka-Kin Ling, Rachel Xiaoyu Wei, Elvis Chun-Sing Chui, Boon Hor Chong, Michael Tim-Yun Ong, Pauline Po-Yee Lui, Patrick Shu-Hang Yung

## Abstract

**Introduction:** Hallux valgus (HV) affects up to 30% of the population. Lapidus surgery with 1st tarsal-metatarsal joint arthrodesis is one of the most common surgical options for HV. Despite its popularity, the current procedure is not without complications. This investigation will be the world’s first Lapidus arthrodesis surgery utilising patient-specific instruments (PSI) as an assistive tool. We hypothesise that the PSI will enhance surgical precision, accelerate fusion rates, decrease non-unions, and reduce the need to use bone grafts.

**Methods and analysis:** This is a single-blinded, parallel-group, randomised controlled trial comparing the outcome of the 3D-Printed PSI Assisted Lapidus Fusion (n=27) vs Conventional Lapidus Fusion (n=27) for HV deformity. Both groups will receive an identical postoperative rehabilitation of protected weight bearing and splinting. Outcomes will include foot function scores, radiological alignment and arthrodesis site assessment with X-ray and High-Resolution Peripheral Quantitative-Computed Tomography, and foot pressure analysis.

**Strengths and Limitations:** *Strengths:*

- This will be the world’s first randomised control trial utilising PSI for hallux valgus surgery.
- Surgeries are done by a team of experienced foot and ankle surgeons.

*Limitations:*

- A single-centre study with a follow-up of only 1 year. However, the authors believe that traditional Lapidus surgery has known term results, and the chief hypothetical benefit of PSI-assisted surgery is faster bony fusion and less surgical complications, which will be evident within 1 year.

## Introduction

Hallux valgus (HV) is one of the most common(1, 2) foot disease among the population; the prevalence could be up to 36%(3). The first ray plays a crucial role in biomechanics for weight bearing and complicated motion during daily activities(4). The instability of the first ray, particularly the first tarsometatarsal joint (TMTJ), is one of the key associations in HV(5-7). Lapidus arthrodesis is a commonly used procedure to attain medial column stability for hallux valgus correction(8).

The most common complication of Lapidus arthrodesis is a high non-union rate(9) which may require revision procedures like bone grafting and revision fixation(10). Inadequate contact of the fusion surfaces due to suboptimal osteotomies lead to delayed fusion and subsequent loss of implant stability. Aside from difficulty getting a perfect osteotomy contact plane, the post-arthrodesis alignment of the 1st metatarsal is also an important factor determining treatment success(11). Optimal 1^st^ metatarsal length is important for the distribution of foot pressure and prevention of postoperative metatarsalgia(12). However, it is difficult to utilise conventional intra-operative 2D fluoroscopy to create a precise cutting plane since hallux valgus is not a simple single-plane deformity but a complex multi-planar deformity(13, 14). Pre-operative planning with 3D CT has been increasingly used in orthopaedics, especially in navigation and robotic-assisted surgery around the spine and knee. However, it is difficult to apply trackers in the foot and ankle region due to the multiple articulations and relatively smaller bones. 3D printing technology has also been applied to orthopaedic surgeries (15) and has allowed the creation of customised patient-specific cutting jigs. The technology is currently applied to high tibial osteotomies (16), fracture fixation (17), tumour resection (18) and spinal surgery(19), with previous studies showing promising results for its application.

In brief, image acquisition, image processing and 3D printing are the three main steps during pre-operation. After acquiring high-quality medical images from CT and processing the images, computeraided design (CAD) will transform the STereoLithography file (STL), which represents the 3D model of the interested objects, into a series of cross-sectional layers. Several methods, including stereolithography apparatus (SLA), fused deposition modelling (FDM), selective laser sintering (SLS) and electron beam melting (EBM), are utilised for printing these objects with the appropriate materials (15). The printed jigs are able to provide a visual and tactile understanding of the anatomical and pathological information of each specific patient. This will greatly help the planning for Lapidus surgery, considering the complex 3D anatomical deformation occurring at the TMTJ.

Duan et al. investigated the availability of 3D-printed customised guides assisting subtalar joint arthrodesis and found that it improved accuracy for drilling the Kirschner wires into the ideal position, reducing the operative time as well as reducing intra-operative radiation(20). The application has also been reported for complex foot reconstruction surgery in patients with Charcot arthropathy(21). The use of 3D-printed jigs in orthopaedics is a relatively new technology. To our best knowledge, no published clinical studies have investigated the outcomes of using 3D-printed jigs in hallux valgus correction surgery.

## Methods and analysis

The study will comply with The Declaration of Helsinki.

### Study Design

This is an open-label, randomised controlled trial where a team of foot and ankle specialists shall be the surgeons. An ITT (intent-to-treat) analysis shall include all randomised patients that not only fit our inclusion and exclusion criteria but gave consent to join this study. From recruitment to outcome measurement timeline please refer to the consort diagram.

### Study Setting

Department of Orthopaedics and Traumatology, Prince of Wales Hospital, Hong Kong

### Objectives and hypothesis

The objective of the current study is to investigate whether a 3D printed patient-specific Lapidus arthrodesis jig will 1) improve bone-to-bone arthrodesis success rates; 2) resume more normal post-operation gait and foot pressure distribution in HV patients. We hypothesise that a patient-specific Lapidus arthrodesis jig will decrease non-union rates and improve clinical outcomes. Measured outcomes include: FAOS score, X-rays, HR pQCT, deformity severity, delayed-union rate, non-union rate and plantar pressure distribution at the relevant designated timeline (refer to Outcomes).

### Sample Size Estimation

Unfortunately, no information was available from previous similar studies which also investigated the clinical outcomes of using 3D printed jigs in Lapidus procedure to correct HV deformity based on Foot and Ankle Outcome Score (FAOS). Therefore, the FAOS Pain Score from a previous survey, which conducted the validation of FAOS for Hallux Valgus, was extracted as the standard deviation (SD) of that in pre-surgery HV patients (SD = 19.4) (22).

When using FAOS to evaluate the improvement of pain in HV, a 15.3-point difference is the smallest score change achievable by an individual and is considered a Minimally Important Change (MIC) (Sierevelt et al., 2016). Using the Normally Distributed Continuous Data-comparing Two Means, a standardised effect size of 0.79 was calculated (Effect size = Mean difference/SD) (23). With a two-sided 5% significance level and 80% power, an estimated sample size in each group of 27 was figured out using G*Power 3.1.

### Allocation

To achieve a lower risk of bias, blocked randomisation will be utilised to form the allocation list for two groups (24). A computer random number generator will be used to select random permuted blocks with a block size of 20 and an equal allocation ratio.

### Participants

Inclusion criteria:

- All hallux valgus patients scheduled for primary Lapidus arthrodesis.
- Hallux Valgus Angle >20°
- 1,2 Inter-metatarsal Angle >9°
- Age >18

Exclusion criteria:

- Disabilities (both physical and mental) which may impair adherence to the rehabilitation.
- Revision surgery
- Concomitantly undergone additional procedures on the same foot (e.g. lesser toe surgery)
- Recent (3 months) use of medications which may influence bone turnover (e.g. chemotherapy, osteoporotic medications)
- Medical comorbidity leading to contraindication for surgery (e.g. active infection, severe uncontrolled diabetes etc.)
- Mentally/physically unable to consent

### Patient and public involvement

Patients who fit both the inclusion and exclusion criteria are briefed with the randomised controlled trial nature of this study. They are informed that all randomisation is done via a computer program, and patients and subsequent doctors they see in outpatient clinics are blinded to the subject group allocation. After the surgery, patients can reflect their own assessment via finishing the FAOS survey (one of the primary outcomes). Results of the study shall be shared with the involved patients via phone if prompted by these participants after this study has ended.

### Intervention

Interventions for each group with sufficient detail to allow replication, including how and when they will be administered. For the experiment group, there are three main steps of the surgery.

#### 1. Surgical Planning

DICOM files will be imported into the Model Intestinal Microflora in Computer Simulation (MIMICS 21.0) 3D image processing software (Materialise, Belgium) for 3D reconstruction. A threshold-based segmentation of the CT images (figure 2) make use of the bone, generating a 3D reconstruction of the medium cuneiform and the 1st metatarsal of the involved foot for surgical simulation. The corresponding surgeon will define the cutting plane on the cuneiform and the metatarsal respectively, which produces the bone segment that will be removed for further fusion. This will help provide electronic data for the design of the computer-aided modelling (CAM) surgical jigs (figure 3).

**Figure 1.**
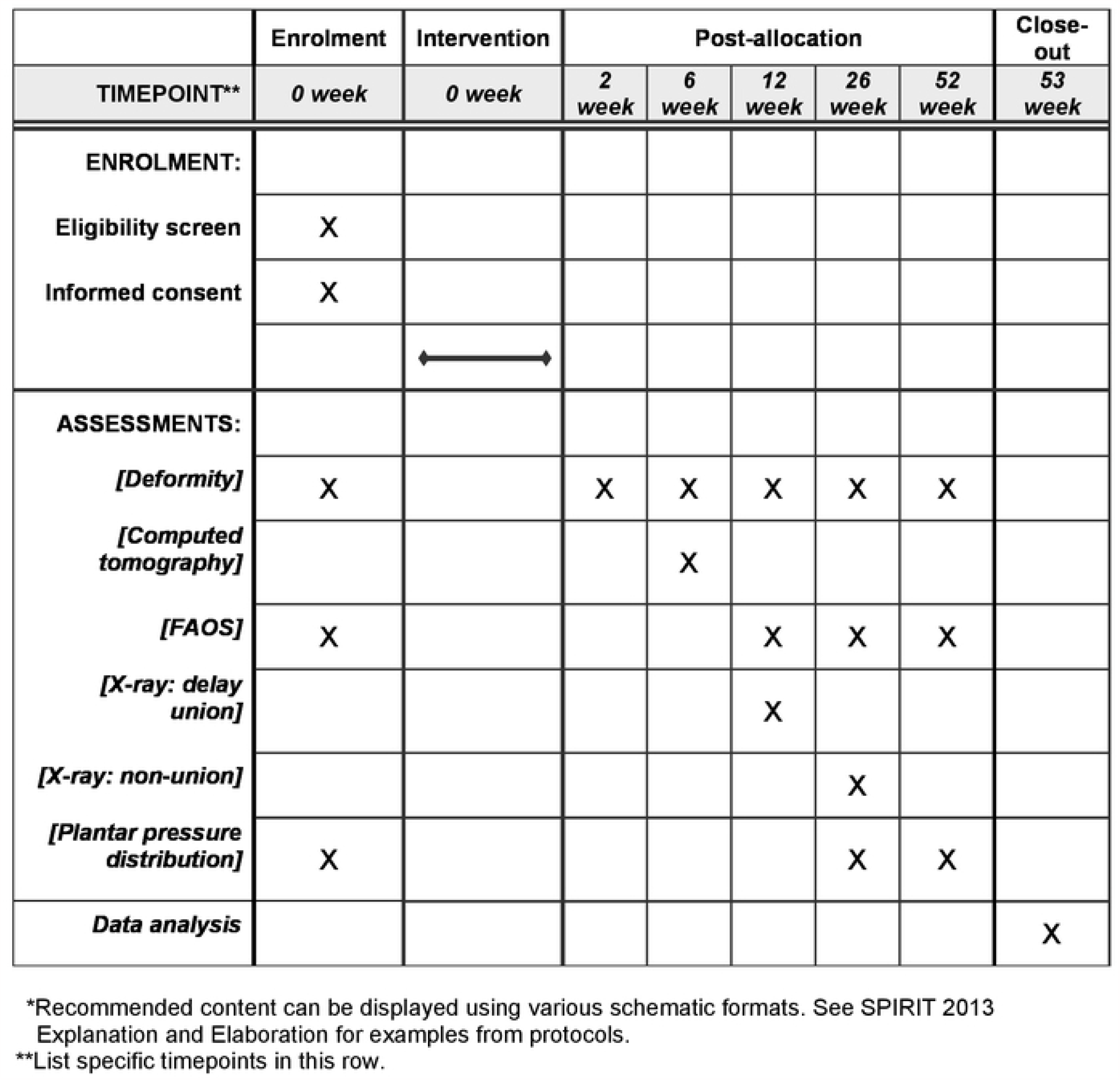
Spirit schedule.

**Figure 2:**
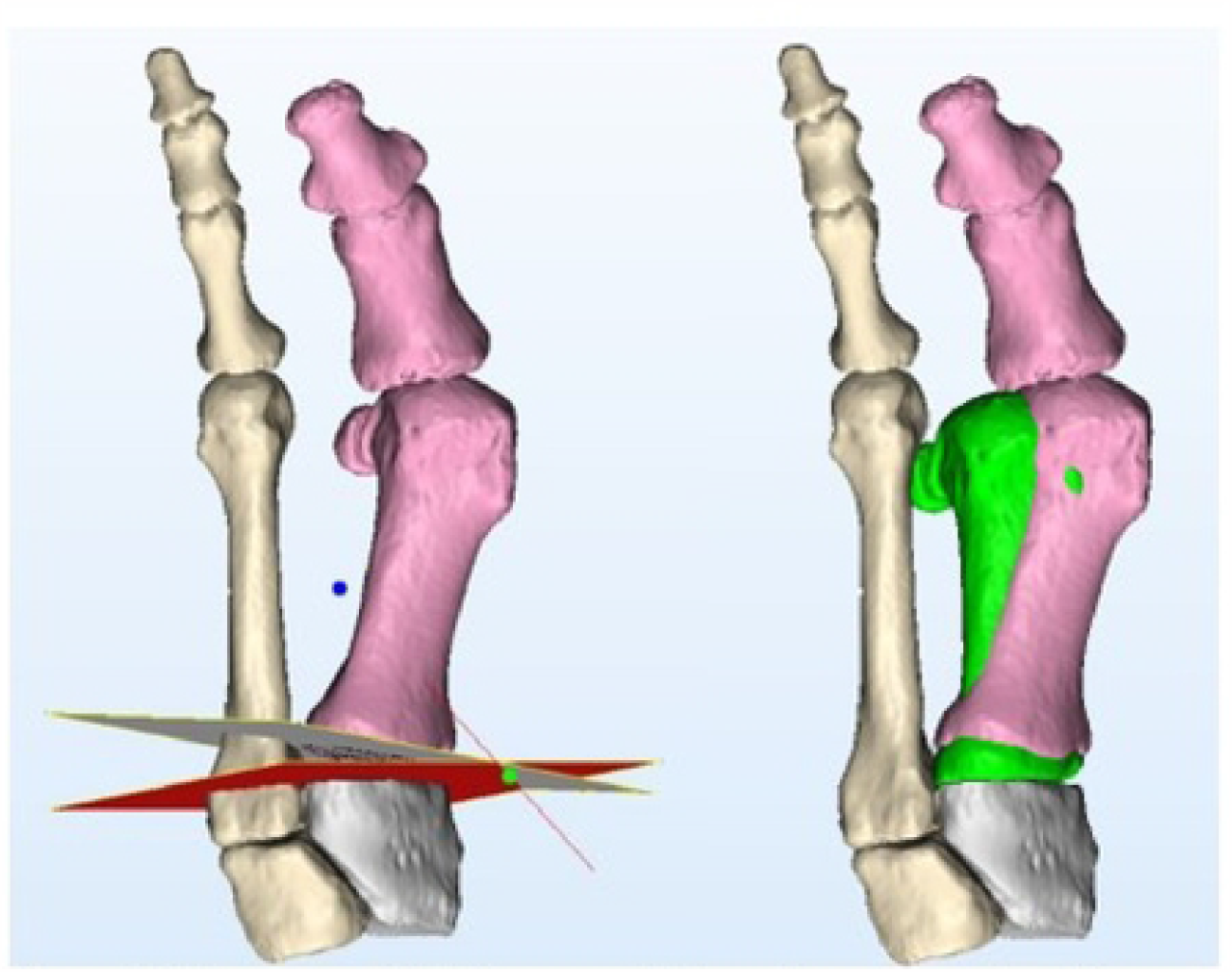
Segmented CT images with cutting plane planning and anticipated final position (green)

**Figure 3:**
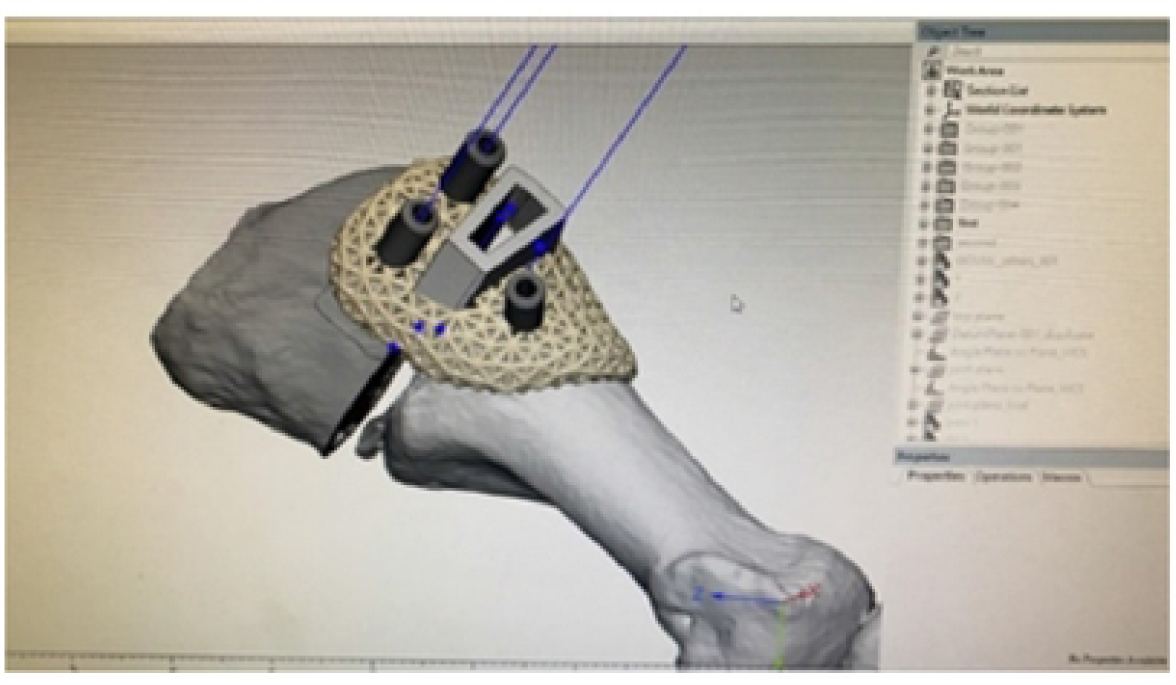
Design of cutting jig

#### 2. Prepare for CAM surgical jigs

The data of the designed jigs were converted into stereolithographic (STL) format and imported into a 3D printer (Fortus 400mc Fused Deposition Modeling system, Stratasys Inc., Eden Prairie, MN) for printing.

#### 3. Lapidus Surgery

After exposing the TMT (tarsometatarsal) joint completely by a medial longitudinal incision and capsulotomy, the custom jig will be set on the surface at the planned bone resection site. The surgeon will hold the jig against the bone by manual pressure. An oscillating saw will then remove the bone segment from the medium cuneiform and the base of the 1st metatarsal according to the jig-based guidance (figure 4). The free remaining bone segment will be adjusted to a normal alignment and be compressed for fusion. The Lapidus arthrodesis will be fixed with two 3.5mm cross-joint headless compression screws. In addition, an arthroscopic assisted lateral soft tissue release with a fluoroscopic assisted bunionectomy will be performed. (1, 25, 26)

**Figure 4:**
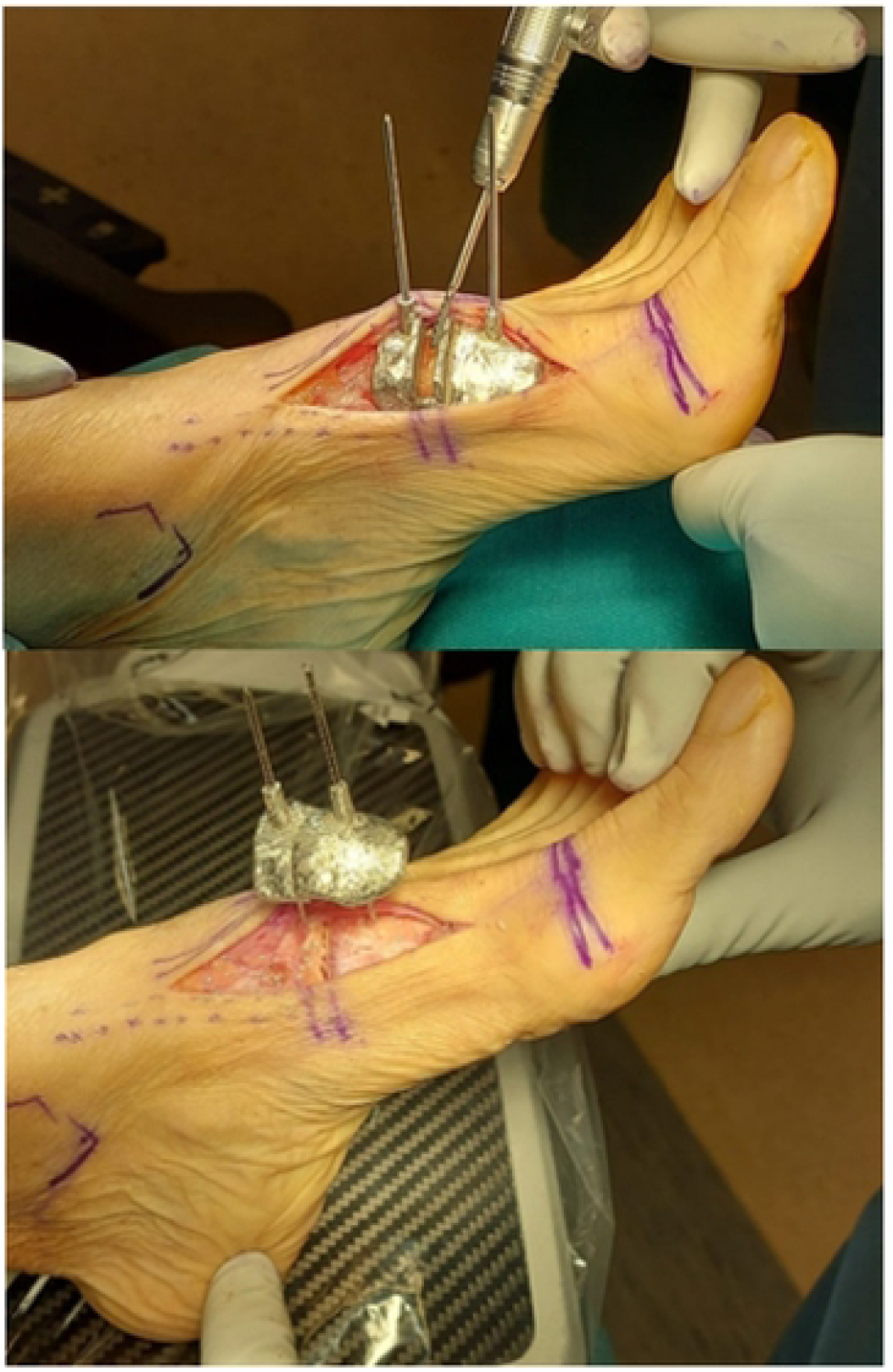
(top) lntraoperative photo of arthrodesis site preparation with a side cutting burr through the”cutting jig”. (bottom) Intraoperative photo showing good compression at the arthrodesis site after application of the “fixation jig”.

For the control group, fluoroscopic assisted open arthrodesis, a fixation with two 3.5mm cross-joint headless compression screws as per routine, and a pairing with an arthroscopic assisted lateral soft tissue release with a fluoroscopic assisted bunionectomy as in the intervention group shall be performed..

Both groups will undergo the same rehabilitation - 2 weeks of non-weight bearing walking(25) followed by 10 weeks of heel walking using a sandal.

##### Outcomes

(Timeline of the outcomes, please refer to consort diagram)

### Primary outcomes

#### 1. Foot and Ankle Outcome Score (FAOS)

FAOS is a reliable and validated patient-reported questionnaire widely used in clinical settings. It consists of five subscales: pain, symptoms, activities of daily living, ability to perform sports and recreational activities and quality of life.(2) The score of each part is recorded on a 0–100 scale, with 100 representing no symptoms. Chinese FAOS has passed all its validity and reliability. (27)

#### 2. X-ray feet

Lateral and dorsal-plantar foot X-rays shall be ordered to investigate if there are any trabeculations crossing the fusion site. Should there be trabeculation extending more than 11/2 of the fusion site, this shall be a successful union.

#### 3. High-resolution peripheral quantitative-Computed Tomography (HR pQCT)

This allows us to visualise the bony micro-architecture at the fusion site and is a more accurate investigation of bone growth evaluation compared to traditional X-rays.(7) With a region of interest at the fusion site, both inner callus and external callus, the volumetric changes in bone mineral density in the area mentioned above shall be further analysed.

### Secondary Outcomes

#### 4. Deformity severity

1,2 intermetatarsal angle (IMA) and hallux valgus angle (HVA) will be measured using dorsal-planar weight-bearing X-rays. By drawing an angle from lines bisecting both the 1^st^ and 2^nd^ metatarsal bone shaft, we will have IMA. On the other hand, by calculating the angle between 1^st^ metatarsal bone shaft and 1^st^ proximal phalanx shaft, we can derive the HVA angle.

#### 5. Delayed-union rate

Delayed union was defined as greater than 50% lucency on either the AP and lateral radiographs or broken hardware at the fusion site at 12 weeks post-surgery. If lucency, sclerosis, or lack of trabeculation extended more than one-half the length of the fusion site on either the A-P or lateral radiograph, a radiographic delayed union was declared (28).

#### 6. Non-union rate

Non-union was defined as greater than 50% lucency on either the AP and lateral radiographs or broken hardware at the fusion site at 26 weeks post-surgery. If lucency, sclerosis, or lack of trabeculation extended more than one-half the length of the fusion site on either the A-P or lateral radiograph, a radiographic non-union was declared (28).

#### 7. Plantar pressure distribution

It has been demonstrated that plantar pressure distribution is highly related to foot biomechanics (29), and plenty of previous work has investigated the change of pressure distribution in HV patients (30, 31). Variation has been found between HV and normal populations. More importantly, this deviation of pressure may lead to transferred pain, namely, metatarsalgia. Considering the potential influence of shortened metatarsal after Lapidus and consequential changed foot biomechanics, the plantar pressure distribution is worthwhile observing in HV patients who receive Lapidus procedure.

The Tekscan Matscan (Tekscan Inc., Boston MA) system will be used to measure the plantar pressure at different anatomical regions during the gait cycle. The sampling frequency of the map will be set as 40 Hz, which is considered sufficient for such data collection (32). Patients will be asked to walk at a self-selected speed across the mat. The two-step method will be adopted, meaning participants will step on the pressure map on the second step (32). The scan will then be masked, and the foot will be divided into eight regions: the hallux, lesser toes, lateral and medial forefoot, lateral and medial midfoot, and lateral and medial hindfoot (33). Peak pressure and impulse in each region will be calculated to see the before-and-after change. Additionally, the centre of pressure excursion index (CPEI) will also be calculated, reflecting the excursion of the centre of pressure. The first and last points of a centre of pressure curve will be connected to construct a line measured in the distal tertile of the foot and normalised by the foot’s width (34).

#### 8. Intraoperative complications

All intra-operative complications will be recorded and documented.

Ethical approval will be sought from the ethical committee from the Chinese University of Hong Kong. At the beginning of each experiment, the research purpose and the experimental procedures will be clearly explained to the participants, and they will be asked to sign a Consent Form before being tested.

### Timeline

#### Baseline assessment

All the patients will come to the clinic and receive the assessment one week before their operation date.

#### Post-operation assessment

All the patients will return for the assessment at 0 weeks, 2 weeks, 6 weeks, 12 weeks, 26 weeks and 52 weeks post-operation. (Refer to consort diagram attached)

### Statistics

For all the primary and second outcomes, an independent t-test will be used to compare the difference between the PSI and the control groups. Repeated measured ANOVA will be used to investigate the changes over time at pre-surgery, 0-week, 12-week, 26-week and 1-year post surgery in the PSI group.

### Data monitoring

A research assistant (both reporting directly to the principal investigator) will be responsible for data collection while the principal investigator will be responsible for data analysis. The principal investigator himself will make decisions to terminate the trial if needed.

### Ethics and dissemination

Research ethics application will be submitted to The Joint Chinese University of Hong Kong – New Territories East Cluster Clinical Research Ethics Committee (The Joint CUHK-NTEC CREC).

### Consent

The principal investigator or co-investigators will obtain informed consent from participants.

### Confidentiality

All collected personal data and medical information relevant to this study about the subjects will be strictly confidential. Subjects will only be identified by a study number and initials in the study database, and no personal identity will be disclosed when study results are being reported and/or published. The data can be accessed only by the principal investigator, co-investigators and the research assistant(s) in charge.

## Data Availability

No datasets were generated or analysed during the current study. All relevant data from this study will be made available upon study completion.

## Author’s contributions

SKKL is the main contributor to the design, surgery and analysis of the data. RXW, ECSC, BHC, PPYL, MTYO helped design and engineer the PSI implant as well as draft the proposal. PSHY will oversee the study.

## Funding statement

This work has been funded by the Health and Medical Research Fund, Health Bureau, the Government of the Hong Kong SAR (Ref 09202926).

## Declaration of interests

The authors declare that they have no conflict of interest.

## Notes

### Competing Interest Statement

The authors have declared no competing interest.

### Clinical Trial

ClinicalTrials.gov Identifier: NCT05602844

### Funding Statement

Yes

### Author Declarations

The Joint CUHK-NTEC CREC Ref 2020.633-T has granted ethics approval for this proposed study.

## Reference

1. Ling SKK, Lui TH. Arthroscopic-Assisted Correction of Hallux Valgus Deformity. Cham: Springer International Publishing; 2016. p. 803–9.

2. Ling SKK, Lui TH. Endoscopy-Assisted Hallux Valgus Correction Provides Sustainable Long-Term >10-Year Outcomes. Arthroscopy: The Journal of Arthroscopic & Related Surgery. 2018;34(6):1958–63.

3. Coughlin MJ, Jones CP. Hallux Valgus: Demographics, Etiology, and Radiographic Assessment. Foot & Ankle International. 2007;28(7):759–77.

4. Rush SM, Christensen JC, Johnson CH. Biomechanics of the first ray. Part II: Metatarsus primus varus as a cause of hypermobility. A three-dimensional kinematic analysis in a cadaver model. The Journal of Foot and Ankle Surgery. 2000;39(2):68–77.

5. Coughlin MJ, Jones CP. Hallux Valgus and First Ray Mobility: A Prospective Study. JBJS. 2007;89(9):1887–98.

6. Dayton P, Kauwe M, Feilmeier M. Is Our Current Paradigm for Evaluation and Management of the Bunion Deformity Flawed? A Discussion of Procedure Philosophy Relative to Anatomy. The Journal of Foot and Ankle Surgery. 2015;54(1):102–11.

7. Wei RX, Ko VM-C, Chui EC-S, Fu BS-C, Hung VW-Y, Yung PS-H, et al. Investigation on the site of coronal deformities in Hallux valgus. Sci Rep [Internet]. 2023 2023/02; 13(1):[1815 p.]. Available from: http://europepmc.org/abstract/MED/36725901

8. Faber FWM, Mulder PGH, Verhaar JAN. Role of First Ray Hypermobility in the Outcome of the Hohmann and the Lapidus Procedure: A Prospective, Randomized Trial Involving One Hundred and One Feet. JBJS. 2004;86(3):486–95.

9. Buddecke DE, Reese ER, Prusa ER. Revision of Malaligned Lapidus and Nonunited Lapidus. Clinics in Podiatric Medicine and Surgery. 2020;37(3):505–20.

10. McInnes BD, Bouché RT. Critical evaluation of the modified lapidus procedure. The Journal of Foot and Ankle Surgery. 2001;40(2):71–90.

11. Petratos DV, Anastasopoulos JN, Plakogiannis CV, Matsinos GS. Correction of adolescent hallux valgus by proximal crescentic osteotomy of the first metatarsal. Acta Orthopædica Belgica. 2008;74(4):496–502.

12. Nakagawa S, Fukushi J-i, Nakagawa T, Mizu-Uchi H, Iwamoto Y. Association of metatarsalgia after hallux valgus correction with relative first metatarsal length. Foot & Ankle International. 2016;37(6):582–8.

13. Wagner P, Wagner E. Role of coronal plane malalignment in hallux valgus correction. Foot and Ankle Clinics. 2020;25(1):69–77.

14. Won H-J, Oh C-S. Variations of the plantar tarsometatarsal ligaments. Clin Anat. 2019;32(5):699–705.

15. Rengier F, Mehndiratta A, von Tengg-Kobligk H, Zechmann CM, Unterhinninghofen R, Kauczor HU, et al. 3D printing based on imaging data: review of medical applications. International Journal of Computer Assisted Radiology and Surgery. 2010;5(4):335–41.

16. Jones GG, Jaere M, Clarke S, Cobb J. 3D printing and high tibial osteotomy. EFORT Open Rev. 2018;3(5):254–9.

17. Kim HN, Liu XN, Noh KC. Use of a real-size 3D-printed model as a preoperative and intraoperative tool for minimally invasive plating of comminuted midshaft clavicle fractures. Journal of Orthopaedic Surgery and Research. 2015;10(1):91.

18. Helguero CG, Kao I, Komatsu DE, Shaikh S, Hansen D, Franco J, et al. Improving the accuracy of wide resection of bone tumors and enhancing implant fit: A cadaveric study. Journal of Orthopaedics. 2015;12:S188–S94.

19. Cai H, Liu Z, Wei F, Yu M, Xu N, Li Z. 3D Printing in Spine Surgery. Adv Exp Med Biol. 2018;1093:345–59.

20. Duan X, Fan HQ, Wang FY, He P, Yang L. Application of 3D-printed Customized Guides in Subtalar Joint Arthrodesis. Orthop Surg. 2019;11(3):405–13.

21. Giovinco NA, Dunn SP, Dowling L, Smith C, Trowell L, Ruch JA, et al. A Novel Combination of Printed 3-Dimensional Anatomic Templates and Computer-assisted Surgical Simulation for Virtual Preoperative Planning in Charcot Foot Reconstruction. The Journal of Foot and Ankle Surgery. 2012;51(3):387–93.

22. Chen L, Lyman S, Do H, Karlsson J, Adam SP, Young E, et al. Validation of Foot and Ankle Outcome Score for Hallux Valgus. Foot & Ankle International. 2012;33(12):1145–55.

23. Sierevelt IN, van Eekeren IC, Haverkamp D, Reilingh ML, Terwee CB, Kerkhoffs GM. Evaluation of the Dutch version of the Foot and Ankle Outcome Score (FAOS): Responsiveness and Minimally Important Change. Knee Surg Sports Traumatol Arthrosc. 2016;24(4):1339–47.

24. Schulz KF, Grimes DA. Generation of allocation sequences in randomised trials: chance, not choice. The Lancet. 2002;359(9305):515–9.

25. Ling SKK, Wu Y-M, Li C, Lui TH, Yung PS-H. Randomised control trial on the optimal duration of non-weight-bearing walking after hallux valgus surgery. Journal of Orthopaedic Translation. 2020;23:61–6.

26. Ling SKK, Lui TH, Yung PSH. Arthroscopic Lateral Soft Tissue Release for Hallux Valgus. The Journal of Foot and Ankle Surgery. 2020;59(1):210–2.

27. Ling SKK, Chan V, Ho K, Ling F, Lui TH. Reliability and validity analysis of the open-source Chinese Foot and Ankle Outcome Score (FAOS). Foot (Edinb). 2018;35:48–51.

28. Mani SB, Lloyd EW, MacMahon A, Roberts MM, Levine DS, Ellis SJ. Modified Lapidus Procedure with Joint Compression, Meticulous Surface Preparation, and Shear-Strain-Relieved Bone Graft Yields Low Nonunion Rate. HSS J. 2015;11(3):243–8.

29. Stolwijk NM, Keijsers NLW, Pasma JH, Nanhoe-Mahabier W, Duysens J, Louwerens JWK. Treatment of metatarsalgia based on claw toe deformity through soft tissue release of the metatarsophalangeal joint and resection of the proximal interphalangeal joint: Evaluation based on foot kinematics and plantar pressure distribution. Foot and Ankle Surgery. 2020;26(7):755–62.

30. Hida T, Okuda R, Yasuda T, Jotoku T, Shima H, Neo M. Comparison of plantar pressure distribution in patients with hallux valgus and healthy matched controls. Journal of Orthopaedic Science. 2017;22(6):1054–9.

31. Martínez-Nova A, Sánchez-Rodríguez R Fau - Pérez-Soriano P, Pérez-Soriano P Fau - Llana-Belloch S, Llana-Belloch S Fau - Leal-Muro A, Leal-Muro A Fau - Pedrera-Zamorano JD, Pedrera-Zamorano JD. Plantar pressures determinants in mild Hallux Valgus. Gait & Posture. 2010;32(3):425–7.

32. Zammit GV, Menz HB, Munteanu SE. Reliability of the TekScan MatScan®system for the measurement of plantar forces and pressures during barefoot level walking in healthy adults. Journal of Foot and Ankle Research. 2010;3(1):11.

33. Galica AM, Hagedorn TJ, Dufour AB, Riskowski JL, Hillstrom HJ, Casey VA, et al. Hallux valgus and plantar pressure loading: the Framingham foot study. Journal of Foot and Ankle Research. 2013;6(1):42.

34. Song J, Hillstrom HJ, Secord D, Levitt J. Foot type biomechanics. comparison of planus and rectus foot types. J Am Podiatr Med Assoc. 1996;86(1):16–23.

